# Diffusion tensor imaging of white matter microstructure in chronic pain: a tract-based spatial statistics study and a systematic review

**DOI:** 10.1101/2020.04.16.20068197

**Authors:** Dominique Christopher Gronemann, Katharina Koch, Carsten Bantel, Peter Sörös

## Abstract

The pathophysiology of many chronic pain disorders is far from evident. MR imaging studies provided initial data indicating chronic pain might lead to changes in brain structure and function. These changes may contribute to cognitive and emotional impairment and maybe even to the chronification of pain. However, the evidence for pain-related changes in gray and white matter is inconclusive so far. Hence we investigated potential changes of white matter microstructure in 34 adults with chronic noncancer pain (> 1 year) and 34 sex- and age-matched healthy individuals using diffusion tensor imaging (DTI). Whole-brain tract-based spatial statistics (TBSS) analyses of fractional anisotropy, mode of diffusivity, mean diffusivity, axial diffusivity, and radial diffusivity did not show significant differences after correction for multiple comparisons. The volumes of subdivisons of the corpus callosum were not significantly different either. We also performed a systematic review of the existing literature on white matter microstructure in patients with chronic pain following PRISMA guidelines. We identified 53 eligible studies on DTI in chronic pain. These studies demonstrated conflicting results regarding the direction and location of white matter changes across all diagnoses and within major diagnostic subcategories. We propose that two factors primarily contribute to this low reproducibility, small sample size and the use of potentially unreliable DTI parameters derived from the single-tensor model.

## Introduction

Chronic pain is a major medical, social, and economical problem, causing a significant individual and societal burden worldwide (1, 2). Chronic pain is often associated with cognitive (3–5) and emotional (6, 7) disturbances and also has serious consequences for social relationships, work, and the individual’s family (8). The prevalence of moderate-to-severe chronic noncancer pain in the adult population ranges between 10% and 20% in developed countries (9, 10). Accordingly, the financial costs of chronic pain are substantial, both for the affected individual and for society in general (11, 12). While the notion that chronic pain is the result of several diverse mechanisms, including neuroscientific, psychological, and social factors, according to Engel’s biopsychosocial model (13, 14), has gained wide support, the exact etiologies of most forms of chronic pain are far from established.

Regarding the neural mechanisms of chronic pain, numerous studies have suggested that pain reshapes the brain’s structure and function and that pain-induced neural reorganization contributes to the chronification of pain (15–17). In a seminal structural MRI study, Apkarian et al. (18) were able to demonstrate gray matter loss in a group of patients with chronic back pain compared to healthy controls. Since then, various studies on gray matter changes in chronic pain have been published with conflicting results regarding the location and direction (decrease vs. increase) of changes. Coordinate-based quantitative meta-analyses have been performed to condense these findings, e.g. (19, 20). These meta-analyses identified areas of decreased and increased gray matter in patients with chronic pain. Importantly, areas of gray matter alterations included not only parts of the nociceptive system, such as the primary somatosensory cortex, the thalamus, and the insula, but also areas outside the core nociceptive system, such as the frontal lobe. The mechanisms that lead to gray matter changes in chronic pain are unknown so far. However, structural MR images of patients with chronic pain did not provide evidence for global neurodegeneration or premature aging of the brain (21).

In the meantime, it has become increasingly clear that also white matter fiber tracts play a crucial role in neural function. Disruption of fiber tracts is e.g. associated with global cognitive dysfunction as well as a decline of executive functions and verbal memory (22). Soon after the initial studies on gray matter researchers started to investigate potential changes of white matter in patients with chronic pain, including DaSilva et al. (23), Lutz et al. (24), and Sundgren et al. (25). While white matter lesions can be identified on conventional T1- or T2-weighted images, the tissue structure of normal-appearing white matter may be estimated with diffusion tensor imaging (DTI) (26). DTI allows investigating the macrostructure (e.g., the thickness and fiber density of major white matter fiber bundles) and microstructure of white matter (27).

Using DTI, changes in white matter microstrucure were found in patients with various pain disorders, including musculoskeletal pain (28, 29), fibromyalgia (30), trigeminal neuralgia (31–33), primary dysmenorrhea (34–36), and chronic neuropathic pain following spinal cord injury (37, 38). Impaired white matter structure in chronic pain may be of considerable clinical importance because white matter disorder may be associated with cognitive deficits and may predict transition to chronic pain (39). Of note, not all studies found changes in white matter microstructure in chronic pain; several studies did not detect differences between patients and healthy controls, including studies by Ceko et al. (40), Neeb et al. (41), and Petrusi ć et al. (42).

For patients with chronic low back pain, two systematic reviews are available, presenting findings of several MR imaging techniques, including DTI (43, 44). For patients with chronic musculoskeletal pain, another systematic review on structural and functional MR imaging of the brain, including DTI, has been published (45). To the best of our knowledge, there is no systematic review synthesizing all available studies on white matter microstructure across all forms of chronic pain.

The first aim of the present study was to investigate white matter microstructure and the volume of corpus callosum subdivisions. As part of a bigger project, DTI was performed in the ChroPain1 study, a cross-sectional case-control study, including 34 patients with chronic noncancer pain and 34 individually age- and sex-matched healthy controls. To assess white matter microstructure, whole-brain tract-based spatial statistics (TBSS) analyses of fractional anisotropy (FA), mode of diffusivity (MO), mean diffusivity (MD), axial diffusivity (AD), and radial diffusivity (RD) were done. TBSS was used to compare DTI diffusivity parameters between patients and controls and to evaluate, within the chronic pain group, potential associations between diffusivity parameters and pain duration and pain intensity. TBSS has been performed in numerous DTI studies and is considered a standard approach for voxel-based analysis of DTI data (46). Based on previous reports (28, 29), we hypothesized that patients with chronic pain, on average, demonstrate decreased FA in white matter fiber tracts, such as corpus callosum, cingulum, and in the frontal white matter. Several studies found changes in white matter microstructure in the corpus callosum of patients with chronic pain, e.g (30). We, therefore, hypothesized that the macroanatomy of the corpus callosum might also be affected in chronic pain.

The second aim of the present study was to perform a systematic review of the existing literature on white matter microstructure in patients with chronic pain based on PRISMA guidelines (47). The research question of this systematic review was: Are there statistically significant differences of white matter DTI parameters between groups of patients with chronic (> 3 months) pain and healthy controls?

## Results

## Diffusion tensor imaging in the ChroPain1 study

### Description of clinical characteristics

We included 42 patients with chronic pain and 42 healthy controls. Due to missing MRI measurements and missing matching partners, we excluded 8 participants in each group. The demographic, clinical, and neuropsychological characteristics of the chronic pain group (n = 34) and the individually matched healthy control group (n = 34) are presented in **Table 1**. Formal education and verbal IQ, as assessed by a vocabulary test, were significantly lower in chronic pain patients (detailed data on spatial-numeric processing and other cognitive functions in this sample have been presented by Spindler et al. (4)). Although we tried to match for formal education, we were unable to recruit sufficient numbers of healthy controls with lower secondary education. We thus decided not to include formal education in the matching process to avoid the loss of a considerable number of participants.

**Table 1.** Demographic, clinical, and neuropsychological data of healthy controls and patients with chronic pain. Data are presented as mean ± standard deviation. ^1^Eleven-point numerical rating scale; 0: no pain, 10: worst pain imaginable. ^2^lower secondary: *Hauptschulabschluss*; secondary: *Realschulabschluss*; high school diploma: *Abitur*; university degree: *Universitätsabschluss*. ^3^Wortschatztest (WST). ^4^Allgemeine Depressionsskala (ADS-K)

|  | Controls<br>n = 34 | Chronic pain<br>n = 34 | p |
| --- | --- | --- | --- |
| Age (yrs) | 54 ± 8 | 54 ± 8 | 0.965 (t-test) |
| Sex |  |  |  |
| Women, n (%) | 25 (74%) | 25 (74%) | — |
| Men, n (%) | 9 (26%) | 9 (26%) | — |
| Pain duration (yrs.) | — | 17 ± 11 (1-50) | — |
| Pain intensity<sup>1</sup> | — | 6 ± 2 (3-9) | — |
| Handedness |  |  |  |
| Right, n | 32 | 29 | 0.149 (χ<sup>2</sup>) |
| Right converted, n | 1 | 5 |  |
| Left, n | 1 | 0 |  |
| Education<sup>2</sup> |  |  |  |
| Lower secondary, n | 1 | 15 | 0.001 (χ<sup>2</sup>) |
| Secondary, n | 15 | 10 |  |
| High school diploma, n | 6 | 3 |  |
| University degree, n | 12 | 6 |  |
| Verbal IQ<sup>3</sup> | 106 ± 9 | 97 ± 9 | > 0.001 (t-test) |
| Depression score<sup>4</sup> | 5 ± 4 | 16 ± 10 | > 0.001 (t-test) |

All participants were Caucasians born and raised in Germany. All patients with chronic pain were seen in a specialized pain outpatient clinic on a regular basis. Primary pain diagnoses were degenerative spinal disease (n = 21), degenerative joint disease (n = 4), fibromyalgia (n = 7), and abdominal pain (n = 2). Patients were treated with non-opioid analgesics (n = 20, 59%), opioid analgesics (n = 14, 41%), glucocorticoids (n = 2, 6%), antidepressants (n = 16, 47%), and antiepileptic drugs (n = 4, 12%). The mean depression score on the German version of the Center for Epidemiological Studies Depression Scale (CES-D) was significantly higher in chronic pain patients.

#### White matter microstructure in chronic pain patients and healthy controls

Whole-brain TBSS of FA, MO, MD, AD, and RD did not result in significant differences between groups after correction for multiple comparisons. **Figure 1A** displays the mean white matter skeleton used for voxel-based TBSS analysis, **Figure 1B** shows voxels in which FA is significantly decreased compared to healthy controls. After correction for multiple comparisons, no significant voxels remained.

**Fig. 1.**
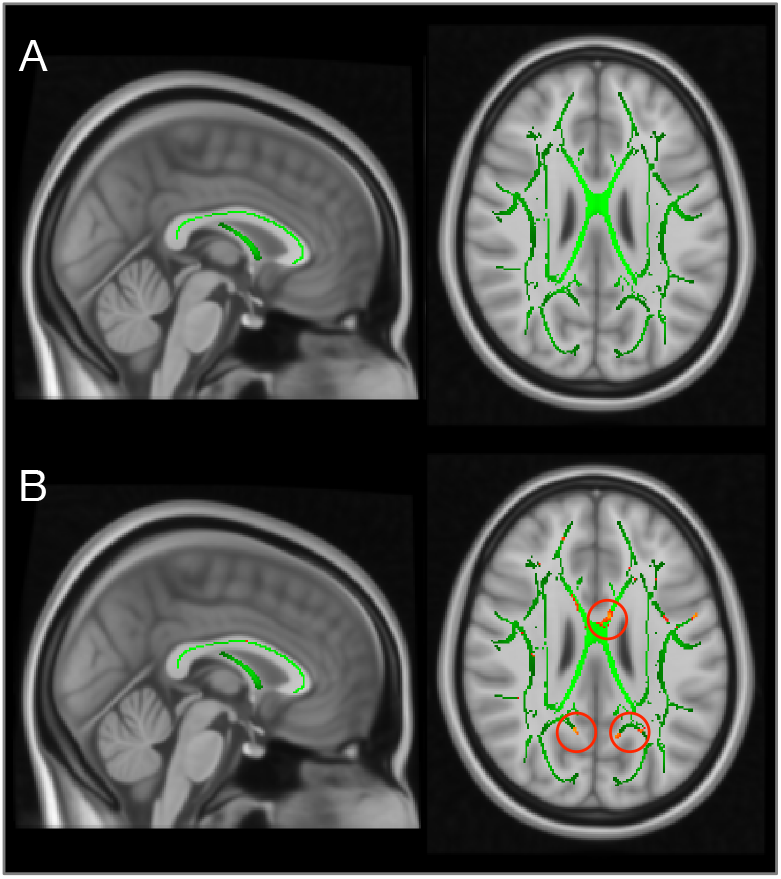
The white matter skeleton used for tract-based spatial statistics (TBSS) is shown in green (**A**). Comparison of fractional anisotropy (FA) between patients with chronic pain and healthy controls (**B**). Voxels in yellow-red represent areas of decreased FA before correction for multiple comparisons.

#### Association between white matter microstructure and pain duration in chronic pain patients

Using whole-brain TBSS, no significant associations between FA, MO, MD, AD, RD, and pain duration were found.

#### Association between white matter microstructure and pain intensity in chronic pain patients

Using whole-brain TBSS, no significant associations between FA, MO, MD, AD, RD, and pain intensity (pain during the last 24 hours before the MRI scan on an 11-point numerical rating scale) were found.

#### Corpus callosum volumetry

Comparing the volumes of five subdivisions of the corpus callosum between patients with chronic pain and healthy controls did not result in statistically significant differences, neither for the original volumes nor for volumes corrected for intracranial volume (Welch’s t-test, p > 0.05).

### Systematic review

#### Study selection

Literature searches in the PubMed and Web of Science Core Collection databases identified 1045 records. The selection process resulted in 53 eligible articles after removing duplicates and applying all inclusion and exclusion criteria (**Table 2**).

**Table 2.**
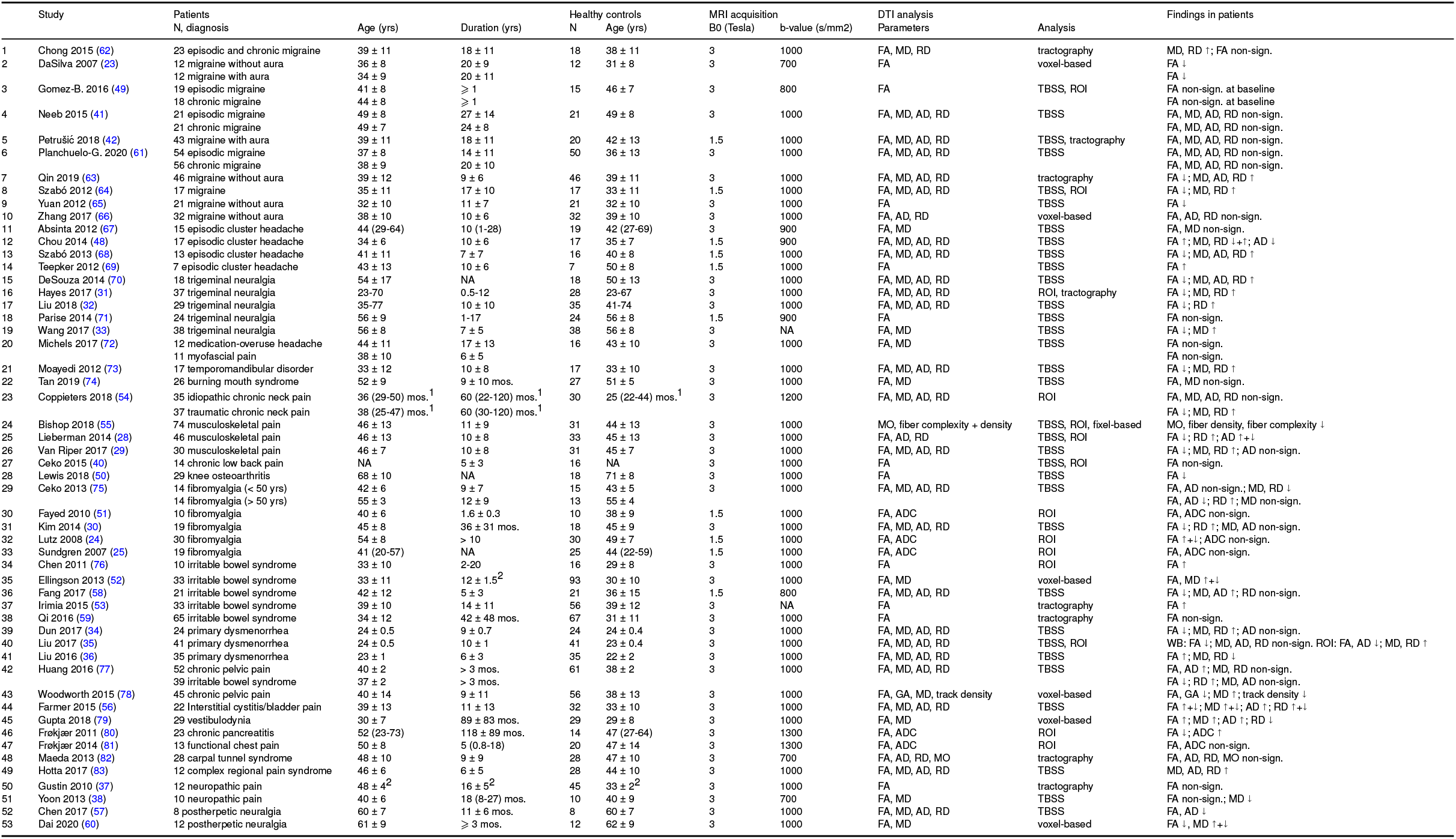
Systematic review: summary of included studies. Data are presented as mean ± standard deviation or mean ± range, unless otherwise stated. FA denotes fractional anisotropy; MD, mean diffusivity; AD, axial diffusivity; RD, radial diffusivity; ADC, apparent diffusion coefficient; GA, generalized anisotropy; NA, not available; TBSS, tract-based spatial statistics; ROI: region-of-interest analysis; WB: whole brain analysis. ^1^median (interquartile range), ^2^mean ± standard error of the mean.

#### Study characteristics

All studies were case-control studies, comparing one or two patient groups with a healthy control group. Four studies were longitudinal (40, 48–50), the remaining studies were cross-sectional.

#### Risk of bias in individual studies

Included studies were very heterogeneous in terms of the specific pain disorder studied, in- and exclusion criteria, and sample size. The majority of studies investigated patients recruited systematically in specialized clinics. The recruitment of healthy controls was less transparent; only a few studies mentioned the recruitment of hospital staff or university students. Several studies mention individual matching (e.g., (41, 51)), but some of these studies recruited unequal sample sizes for patients and controls. The majority of studies performed frequency matching (recruiting a convenience sample of healthy controls with similar age and sex distribution as found in the chronic pain group). Almost all studies presented detailed diagnostic criteria for the patient group and additional inclusion and exclusion criteria for all participants. Two studies did not state clear diagnostic criteria for the patient group (40, 50), another two studies did not state additional inclusion and exclusion criteria (52, 53). Only 16 studies communicated if participants were excluded after enrollment in the study. No study reported that a participant withdrew from the study. One study mentioned a significant difference in age between patient and control group (54), 6 studies did not present statistical results on potential age differences. Five studies mentioned a significant difference in the distribution of men and women (28, 38, 49, 55, 56), 8 did not present statistical results on potential differences in sex. Seven studies also matched for years of education (30, 34, 35, 57–60), in addition to age and sex.

The majority of studies was performed on a 3 Tesla scanner (n = 43), 10 studies used a 1.5 Tesla system. The number of diffusion directions ranged between 6 and 64, the b-values between 700 and 1300 s/mm^2^ (40 studies used 1000 s/mm^2^). No study measured two or more b-values > 0 s/mm^2^).

The sample sizes of all studies are shown in **Figure 2**. The median sample size of the patient group was 23 (minimum 7, maximum 74). Of note, only three studies justified the chosen sample size (50, 53, 61).

**Fig. 2.**
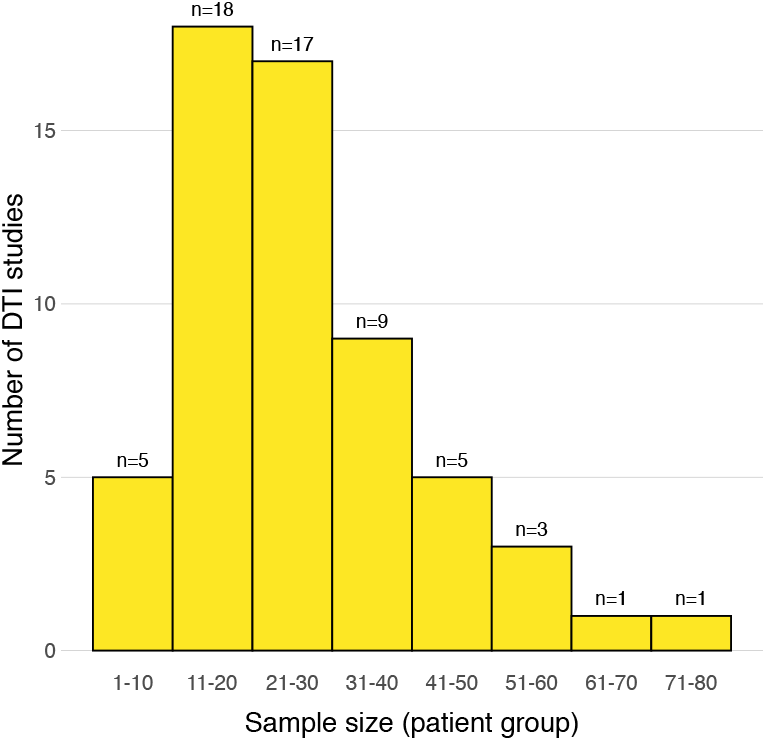
Distribution of sample sizes (patient group only) across all studies included in the systematic review.

The majority of studies used whole-brain voxel-based TBSS. Several studies performed voxel-based comparisons with different software tools or tractography-based analyses.

One study also presented the results of a fixel-based analysis (55) (a fixel represents the different fiber populations within a single voxel (84)).

#### Synthesis of results

The results of individual studies are summarized in **Table 2**. The findings presented in the table list comparisons of patient group(s) with the healthy control group. The results of subgroup analyses (e.g. (42)), comparisons between patient groups (e.g. (61) or correlations of DTI parameters with clinical characteristics are not shown.

Considering all studies on white matter microstructure in chronic pain, the results of the DTI parameters are inconclusive. FA, determined in all studies, MD, AD, and RD are found increased, decreased, and unchanged in patients vs. controls. When separating studies into the major diagnoses, such as migraine, cluster headache, musculoskeletal pain, or fibromyalgia, similarly conflicting effects were seen. However, in two studies on chronic pelvic pain, FA was decreased compared to controls (77, 78). In two studies on neuropathic pain after spinal cord injury, FA was not significantly different compared to controls (37, 38).

## Discussion

The present case-control study investigated white matter microstructure and corpus callosum morphometry in 34 patients with chronic noncancer pain and 34 healthy controls, age- and sex-matched on an individual basis. Whole-brain tract-based spatial statistics (TBSS) analyses of fractional anisotropy (FA), mode of diffusivity (MO), mean diffusivity (MD), axial diffusivity (AD), and radial diffusivity (RD), as well as corpus callosum segmentation, did not reveal statistically significant differences between groups. The aim of the systematic review was to summarize potential differences in white matter microstructure between patients with chronic pain and healthy controls. The results of our review were conflicting and demonstrated a low reproducibility of white matter changes, across all studies on chronic pain and within major diagnostic categories. We propose two main reasons for these conflicting results, (1) insufficient sample sizes, resulting in underpowered studies and (2) the use of an oversimplified diffusion tensor model, resulting in unreliable diffusion parameters.

A major methodological concern of neuroimaging studies is a sufficient sample size (85). Studies with small sample sizes and, consequently, low power are at risk of failing to detect true effects, exaggerating effect sizes, and increasing the false report probability (86–88). As MR scan time is expensive and, for clinical studies, recruiting patients fulfilling strict inclusion criteria is difficult and time-consuming, most neuroimaging studies investigate small samples. This has been shown for functional and structural (T1-weighted) MR studies (89, 90). To our knowledge, there is no systematic assessment of sample sizes in DTI studies.

Heiervang et al. (91) used probabilistic tractography in healthy individuals to define the cingulum bundle, pyramidal tracts, optic radiations, and genu of the corpus callosum and determined mean FA and MD along the tracts. Based on these results, the required sample sizes were calculated to detect reductions in mean tract FA and mean MD with a one-tailed significance level of 0.05, power of 0.8, and equal sample sizes for patients and controls. For a reduction in FA and an effect size of 2%, between 31 and 221 participants per group were required. For a reduction in MD and an effect size of 2%, between 9 and 159 participants per group were required. Unfortunately, we were unable to find effect sizes for DTI studies on patients with chronic pain. We speculate that many of the studies included in our systematic review (and maybe even our ChroPain1 study) were underpowered. The ChroPain1 study and almost all studies summarized in the systematic review used the diffusion tensor model to determine markers of diffusivity for each voxel of the brain. This method gained huge popularity, and FA was frequently interpreted as representing “white matter integrity” (30, 62, 68). However, FA expresses the degree of anisotropy, or directionality of diffusion, in an entire voxel. FA and other DTI parameters are greatly influenced by different factors, such as the MRI hardware and sequences used, technical and biological artifacts, and local tissue properties, such as axonal density, axonal diameter, and degree of myelination (92–94) Another major confound to diffusion tensor estimation is the existence of crossing fibers (or fiber bundles in different directions) in a voxel (84, 94). It has been estimated that up to 90% of all white matter voxels contain crossing fibers (95). Given the aforementioned methodological limitations, our expectation was nevertheless to detect statistically significant differences between the patient and control group in the ChroPain1 study because of the precise individual matching and the high-end MR scanner used with a 64-channel head/neck receive-array coil. One limitation of our study is that we only acquired 27 axial slices to shorten the measurement time and to reduce head motion during the DTI scan (96). As a result, the cerebellum was not covered; potential differences in white matter cerebellar tracts could not be investigated. Another limitation of our study is that we only acquired diffusion images at b-values of 0 s/mm^2^ and 1500 s/mm^2^. Thus, advanced analysis techniques, such as diffusion kurtosis imaging (97) or fixel-based analysis (84) were not possible.

For future studies on potential changes of white matter microstructure in patients with chronic pain we recommend: (1) Investigation of larger sample sizes; a statistical power analysis with G*Power^1^, assuming a two-tailed independent samples t-test with a (hypothetical) medium effect size of d = 0.5 and 95% power, resulted in necessary sample sizes of 105 patients and 105 controls. (2) Exact and individual matching regarding age and sex; cross-sectional and longitudinal studies indicate that healthy aging is associated with a decrease of FA in several brain regions (98). (3) Use of advanced imaging techniques, such as diffusion kurtosis imaging (97) and fixel-based analysis (84) (implemented in MRtrix3 (99)), to overcome the limitations of the diffusion tensor model and its most popular metric, fractional anisotropy.

## Methods

### Diffusion tensor imaging in the ChroPain1 study

#### Participants

This investigation is part of the ChroPain1 study, focusing on the neuropsychological (4) and neural changes (21) associated with chronic pain. Data of 34 patients with chronic pain and 34 healthy controls were analyzed. Patients were recruited from the Pain Outpatient Clinic, University Clinic of Anesthesiology, Critical Care, Emergency Medicine, and Pain Management, Klinikum Oldenburg in Oldenburg, Germany. Additional patients were found through advertisements in the local daily newspaper. For each patient with chronic pain, a sex- and age-matched (± 5 years) pain-free control participant was recruited (individual or pair matching (100), also known as the head-by-head method of recruitment). Healthy controls were identified with the help of the local newspaper, the University of Oldenburg’s web page, flyers, and personal communication.

Following the Classification of Chronic Pain suggested by the International Association for the Study of Pain (101), we used a purely temporal definition of chronic pain. The International Association for the Study of Pain defines chronic pain as pain that lasts or recurs for > 3 months (101). In this study, we extended the minimum duration of pain to 12 months, because we hypothesized that white matter changes will be more recognizable in patients with longer pain duration. The inclusion criterion for the pain group was thus pain on most days of a month for); ⩾ 12 months of mild to severe intensity. This was established by reviewing clinical charts and through patient interviews.

Exclusion criteria for the chronic pain and the healthy control group were as follows: neurological disorders (such as dementia, Parkinson’s disease, stroke, epilepsy, multiple sclerosis, traumatic brain injury, and migraine), psychiatric disorders (such as schizophrenia or major depression), substance abuse, impaired kidney or liver function, and cancer. The ChroPain1 study was approved by the Medical Research Ethics Board, University of Oldenburg, Germany (25/2015). Written informed consent was obtained from all participants before entering the study. Participants received a compensation of 10 €per hour.

#### Calculation of sample size

A formal calculation of the necessary sample size was not possible because we were unable to find effect sizes for tract-based spatial statistics analyses in previous studies on patients with chronic pain.

#### Demographic and clinical data

Participants’ date of birth, sex, handedness, and the highest degree of formal education were recorded. In a structured interview, previous and present conditions that may lead to an exclusion from the study and complete medication records were inquired. Patients were asked to estimate the average pain intensity during the last 24 hours before the MRI examination by using the 11-point numerical rating scale, with 0 representing “no pain” and 10 “worst pain imaginable” (102). Pain duration in years was also noted.

#### Neuropsychological data

To test verbal intelligence, a standardized German vocabulary test (Wortschatztest, WST) was used (103). Participants were required to identify an existing German word within 5 nonwords in each of 42 rows. The number of correct choices was transformed into IQ scores according to the test manual (103) WST results are highly correlated with general intelligence and level of education (104). The Center for Epidemiological Studies Depression Scale (CES-D) (105) in its short German version (Allgemeine Depressionsskala, ADS-K) (106) was used to quantify depressive symptoms. The scale consists of 15 items assessing depressive symptoms during the preceding week. Each item is answered on a 4-point Likert scale: “never or rarely” (<1 day), “sometimes” (1-2 days), “often” (3-4 days), and “always” (5-7 days of the week). The total score ranges from 0 to 45. A score of); ⩾ 18 supports the diagnosis of a clinically relevant depression (106).

#### MR data acquisition

MR images were acquired at 3 Tesla on a MAGNETOM Prisma whole-body scanner (Siemens, Erlangen, Germany) with the XR gradient system (gradient strength = 80 mT/m, gradient rise time = 200 T/m/s on all three gradient axes simultaneously) and a 64-channel head/neck receive-array coil. The scanner is located at the Neuroimaging Unit, School of Medicine and Health Sciences, University of Oldenburg, Germany^2^.

For anatomical brain imaging, Siemens’ 3-dimensional T1-weighted MPRAGE sequence was used with the following parameters: time of repetition (TR): 2000 ms, echo time (TE): 2.41 ms, inversion time (TI): 920 ms, flip angle: 9°, voxel dimensions = 0.7 × 0.7 mm^2^, slice thickness = 0.9 mm, 208 axial slices.

DTI was acquired using a 2-dimensional echo-planar sequence: TR: 3000 ms, TE: 73 ms, voxel dimensions = 2.5 × 2.5 × 2.5 mm^3^, 27 axial slices. Images had an isotropic distribution along 64 directions using a b-value of 1500 s/mm^2^. In addition, 6 volumes with no diffusion weighting were acquired (b = 0 s/mm^2^). T1-weighted and DTI sequences used in-plane acceleration (GRAPPA) with an acceleration factor of 2. Siemens’ pre-scan normalization filter was used in both sequences for on-line compensation of regional signal inhomogeneities.

#### Data analysis

T1-weighted and DTI images were first converted from Siemens DICOM format to compressed NIfTI (.nii.gz) format using dcm2niix^3^. Data conversion with dcm2niix also generates *bvecs* and *bvals* files, containing information about the gradient directions and diffusion weighting applied.

Analysis of T1-weighted and DTI data was carried out using FSL (FMRIB Software Library)^4^ version 6.0.2 (107, 108). DTI analysis was supported by tools and scripts provided by MRItrix3^5^ (99). These analyses were performed under macOS 10.14 Mojave on an Apple iMac.

The segmentation of the corpus callosum was performed by FreeSurfer^6^ version 6.0.0 (109, 110) under Red Hat Enterprise Linux on the high-performance cluster CARL, University of Oldenburg, Germany^7^.

#### Preprocessing of T1-weighted images

T1-weighted images were preprocessed with FSL’s *fsl_anat* script^8^ using the default parameters. This script performs several preprocessing steps, including bias-field correction using FSL’s FAST (FMRIB’s Automated Segmentation Tool) (111) and brain extraction based on transforming a standard-space mask to the input image using FSL’s FNIRT (FMRIB’s Non-Linear Image Registration Tool) (107).

#### Preprocessing of diffusion-weighted images

Preprocessing of DTI data was performed using the MRtrix3 package (99): Thermal noise was reduced using the *dwidenoise* command. This approach performs principal component analysis of the original DTI data and removes noise-only principal components, resulting in signal-to-noise ratio improvements (112, 113). (2) Gibbs ringing artifacts were reduced with the *mrdegibbs* command using the method of local subvoxel-shifts (114). (3) MRtrix3’s *dwipreproc* script was used to perform motion and eddy-current distortion correction (115). The script was used with the *-rpe_none* option because reversed phase-encoding image data were not available, invoking FSL’s *eddy* tool^9^ (115) and using the b = 0 s/mm^2^ volumes as reference. For quality control, all residuals were visually inspected. (4) A whole-brain mask was generated from the preprocessed DTI data set using the *dwi2mask* command.

#### Estimation of diffusion tensors

A single-tensor diffusion tensor model was fitted at each voxel of the preprocessed DTI data using FSL’s *dtifit* program^10^, part of FDT (FMRIB’s Diffusion Toolbox). Using least squares on the log-transformed signal, whole-brain maps of the following parameters were generated (for detailed explanations and equations, see (93, 116)): (1) The first, second, and third eigenvalues (*λ*1, *λ*2, *λ*3), describing the 3 directions of water diffusivity. The first eigenvalue is also referred to as axial diffusivity (AD), representing diffusion parallel to the principal diffusion direction (117), (2) fractional anisotropy (FA), the standard deviation of the 3 eigenvalues divided by their root mean square, (3) mode of anisotropy (MO), the 3^rd^ moment of the tensor (118), (4) mean diffusivity (MD), the average of the 3 eigenvalues and (5) radial diffusivity (RD), the average of the second and third eigenvalues, representing diffusion perpendicular to the principal diffusion direction (117).

#### Tract-based spatial statistics

Voxel-based statistical analysis of DTI parameters was performed using FSL’s TBSS (tract-based spatial statistics)^11^ (119): (1) FA images were preprocessed using the *tbss_1_preproc* command to slightly erode the FA images and zero the end slices. (2) FA data of all participants were aligned into a common space using the FMRIB58_FA_1mm standard-space image^12^ and FSL’s non-linear registration tool FNIRT with the *tbss_2_reg-T* command. (3) Individual participants’ FA images were transformed into MNI152 standard space with the *tbss_3_postreg-S* command. The mean of all individual FA images was then calculated. This mean FA image was thinned to create a mean FA skeleton, which represents all white fiber tracts common to the entire group of participants. (4) The mean FA skeleton was thresholded at 0.2 and all individual FA data were projected onto the mean FA skeleton with the *tbss_4_prestats* command. Statistical comparison was carried out using a binary mask of the FA skeletonized image ensuring that only voxels within each tract were analyzed, preventing partial volume effects.

To perform voxel–based statistics on the skeletonized FA data, FSL’s *randomise* program was used with 10,000 random permutations (120). The statistical threshold was set at p < 0.05, corrected for multiple comparisons by controlling the family-wise error rate and using threshold-free cluster enhancement (TFCE) across all white matter tracts in the whole-brain analysis. TFCE enhances cluster-like features in a statistical image without relying on a pre–defined cluster–forming threshold (121). In addition to FA, TBSS was performed with MO, MD, AD, and RD maps. For each parameter, a non-parametric t-test was used to compare patients with chronic pain and healthy controls. To test if pain duration and pain intensity predicted DTI parameters, linear regressions were performed with pain duration and pain intensity as regressors.

#### Segmentation of the corpus callosum

The volumes of five regions of the corpus callosum were determined by FreeSurfer version 6.0.0 using the *recon-all* command (109, 110). Processing included motion correction, removal of non-brain tissue using a hybrid watershed/surface deformation procedure (122), automated Talairach transformation, and segmentation of subcortical structures (109). The corpus callosum was automatically segmented into the following regions: (1) anterior, (2) midanterior, (3) central, (4) midposterior, and (5) posterior. The volumes of these regions were divided by the estimated total intracranial volume (eTIV) as determined by FreeSurfer (123) to correct for different brain sizes. Original volumetric measurements and corrected values were compared between patients with chronic pain and healthy controls using Welch’s t-test.

#### Systematic review

A systematic review of the available literature on white matter microstructure in patients with chronic pain was performed following the PRISMA guidelines (47).

#### Literature search

We searched the free PubMed search engine provided by NIH’s National Library of Medicine^13^and Clarivate Analytics’ Web of Science Core Collection^14^ through University of Oldenburg’s Library and Information System^15^. For both search engines, search terms were “pain diffusion tensor” and “pain DTI” in all fields and all years. There were no language restrictions. The literature search was initially performed on December 22, 2019 and updated on March 29, 2020. All retrieved bibliographic records were uploaded to the open-access online tool CADIMA^16^ as RIS files. CADIMA supports several steps throughout the systematic review process (124). Here, CADIMA was used for study selection, including duplicate removal, the definition of selection criteria, and screening of records according to the selection criteria at title/abstract and full-text stage. Data extraction was performed off-line after downloading CADIMA’s data extraction sheet containing the bibliographic information of all selected records. The full-text PDF files of all selected papers were imported and managed in DEVONthink 3 for macOS.

#### Study selection

This systematic review includes all original research studies that met the following inclusion and exclusion criteria: Studies included had to (1) be published in a peer-reviewed journal, (2) include a group of patients with chronic or recurrent pain (> 3 months) without neurological or psychiatric disorder affecting the brain (except primary headache disorders), (3) include a group of healthy controls, (4) perform diffusion tensor imaging of the brain in both groups, (5) report a comparison of white matter DTI parameters between chronic pain patients and healthy controls. Studies excluded (1) were published poster abstracts, (2) investigated chronic pain patients with a neurological disorder, such as Parkinson’s disease or stroke, with traumatic brain injury or amputation (to avoid disorder-specific rather than pain-specific effects), (3) and did not present data of white matter microstructure of the brain.

#### Data extraction

For each selected study, we retrieved the following participant data: (1) sample size, main diagnosis, age and pain duration of the patient group(s) and (2) sample size and age of the healthy control group. We also recorded methodological details: (1) field strength of the MRI scanner, (2) number of diffusion directions, (3) b-value, (4) data analysis approach, and (5) main findings of the study relative to the patient group.

#### Risk of bias in individual studies

To assess the methodological quality of individual studies, we modified the Quality Assessment Tool For Quantitative Studies^17^, developed by the Effective Public Health Practice Project (125). To assess selection bias, we recorded the method of patient recruitment and the strictness of diagnostic criteria and additional inclusion and exclusion criteria. We also noted if the study mentioned excluded subjects.

To assess potential confounders, we noted if age and sex were significantly different between patient and healthy control groups. Moreover, we marked if groups were also matched for years of education. Regarding data collection methods, we summarized the technical details of DTI (see the previous section). In addition, we noted sample sizes, justifications of sample sizes, and the use of effect sizes (in addition to p-values). Participant blinding and intervention integrity were not rated. In interventional studies, only baseline results were included in the review.

## Data Availability

The datasets generated during and/or analyzed during the current study are available from the corresponding author on reasonable request.

## ACKNOWLEDGEMENTS

The authors thank Melanie Spindler for assisting with recruitment and data acquisition and Gülsen Yanç and Katharina Grote for helping with MRI acquisition. The study was funded by an intramural grant from the School of Medicine and Health Sciences, University of Oldenburg, to C. Bantel (Forschungspool, 2015- 1). The study was also supported by the Neuroimaging Unit, University of Oldenburg, funded by grants from the German Research Foundation (DFG; 3T MRI INST 184/152-1 FUGG and MEG INST 184/148-1 FUGG).

## Footnotes

1 www.psychologie.hhu.de/arbeitsgruppen/allgemeine-psychologie-undarbeitspsychologie/gpower.html

2 uol.de/en/medicine/biomedicum/neuroimaging-unit

3 github.com/rordenlab/dcm2niix

4 fsl.fmrib.ox.ac.uk/fsl/fslwiki/FSL

5 www.mrtrix.org

6 freesurfer.net

7 uol.de/en/school5/sc/high-perfomance-computing/hpc-facilities/carl

8 fsl.fmrib.ox.ac.uk/fsl/fslwiki/fsl_anat

9 fsl.fmrib.ox.ac.uk/fsl/fslwiki/eddy

10 fsl.fmrib.ox.ac.uk/fsl/fslwiki/FDT/UserGuide#DTIFIT

11 fsl.fmrib.ox.ac.uk/fsl/fslwiki/TBSS

12 fsl.fmrib.ox.ac.uk/fsl/fslwiki/FMRIB58_FA

13 www.ncbi.nlm.nih.gov/pubmed

14 www.webofknowledge.com

15 uol.de/en/bis

16 cadima.info

17 www.city.hamilton.on.ca/phcs/EPHPP

